# Immunochromatographic test for the detection of SARS-CoV-2 in saliva

**DOI:** 10.1101/2020.05.20.20107631

**Authors:** Katsuhito Kashiwagi, Yoshikazu Ishii, Kotaro Aoki, Shintaro Yagi, Tadashi Maeda, Taito Miyazaki, Sadako Yoshizawa, Katsumi Aoyagi, Kazuhiro Tateda

## Abstract

We evaluated the rapid immunochromatographic test for SARS-CoV-2 antigen detection using 16 saliva specimens collected from 6 COVID-19 hospitalized patients, and detected N-antigen in 4 of 7 RT-PCR positive specimens. The POCT antigen test using saliva is highly considered to be a game-changer for COVID-19 diagnosis.

## Introduction

COVID-19 caused by SARS-CoV-2 infection is diagnosed by nucleic acid amplification test, as reverse-transcription polymerase chain reaction (RT-PCR), for viral genome RNA in nasopharyngeal swab specimens. The collection procedure of nasopharyngeal specimens poses a risk of secondary infection to health-care workers. The risk could be reduced if saliva specimens could be used for diagnosis of COVID-19. Recently, several reports indicated that SARS-CoV-2 was detected in saliva specimens of equally or higher sensitivity than the nasopharyngeal specimens[1–3].

We recently developed a rapid antigen detection kit, ESPLINE® SARS-CoV-2 test, for detection of the viral N antigens in nasopharyngeal swabs (manuscript in preparation). The ESPLINE test is an enzyme-immunoassay based on immunochromatographic technology using monoclonal antibodies specific to SARS-CoV and CoV-2 N antigens. In this report, we evaluated the saliva specimens for diagnosis of COVID-19 by RT-PCR and the point of care testing (POCT) antigen test.

### Materials and methods

Sets of saliva and nasopharyngeal specimens were collected within several days from 6 COVID-19 patients hospitalized in Omori Hospital, who had been diagnosed by RT-PCR test. Informed consent was obtained from all participants in the study. The study protocol was approved by the Ethics Committee of Faculty of Medicine, Toho University (No. A20028_A20020_A20014_A19099). Saliva was collected by passive drool method according to previous report[4], Ct values were obtained by RT-PCR test using N2 probe according to the manual provided by National Institute of Infectious Diseases[5], SARS-CoV-2 antigen was assessed using ESPLINE kit according to the manufacturer’s instruction (Fujirebio Inc., Tokyo). The procedure is briefly described as follows: specimen collected by the Swab in the kit was treated with the sample treatment solution. Approximately 20 μL of the treated sample was applied to the immunochromatography cassette, and was developed for 30 minutes. After incubation, test and reference lines were visually assessed.

## Results

SARS-CoV-2 RNA was detected in 13 of 16 (81.3%). On the other hand, RT-PCR detected RNA in only 7 of 16 saliva specimens (43.8%). No significant difference in RNA copies of nasopharyngeal specimens between positive and negative RT-PCR results of saliva specimens was found. When 9 of the 13 RT-PCR positive nasopharyngeal specimens were examined, SARS-CoV-2 antigen in 7 specimens (77.8% in sensitivity) was detected by the ESPLINE test. The antigen was detected in 4 of 7 RT-PCR positive saliva specimens (47.8%)(Figure 1). As shown in previous studies of SARS-CoV-2 antigen tests, sensitivity of the antigen seems to be correlated with RNA copies concentrations in the specimens^5^ (Manuscript in preparation).

**Figure 1.**
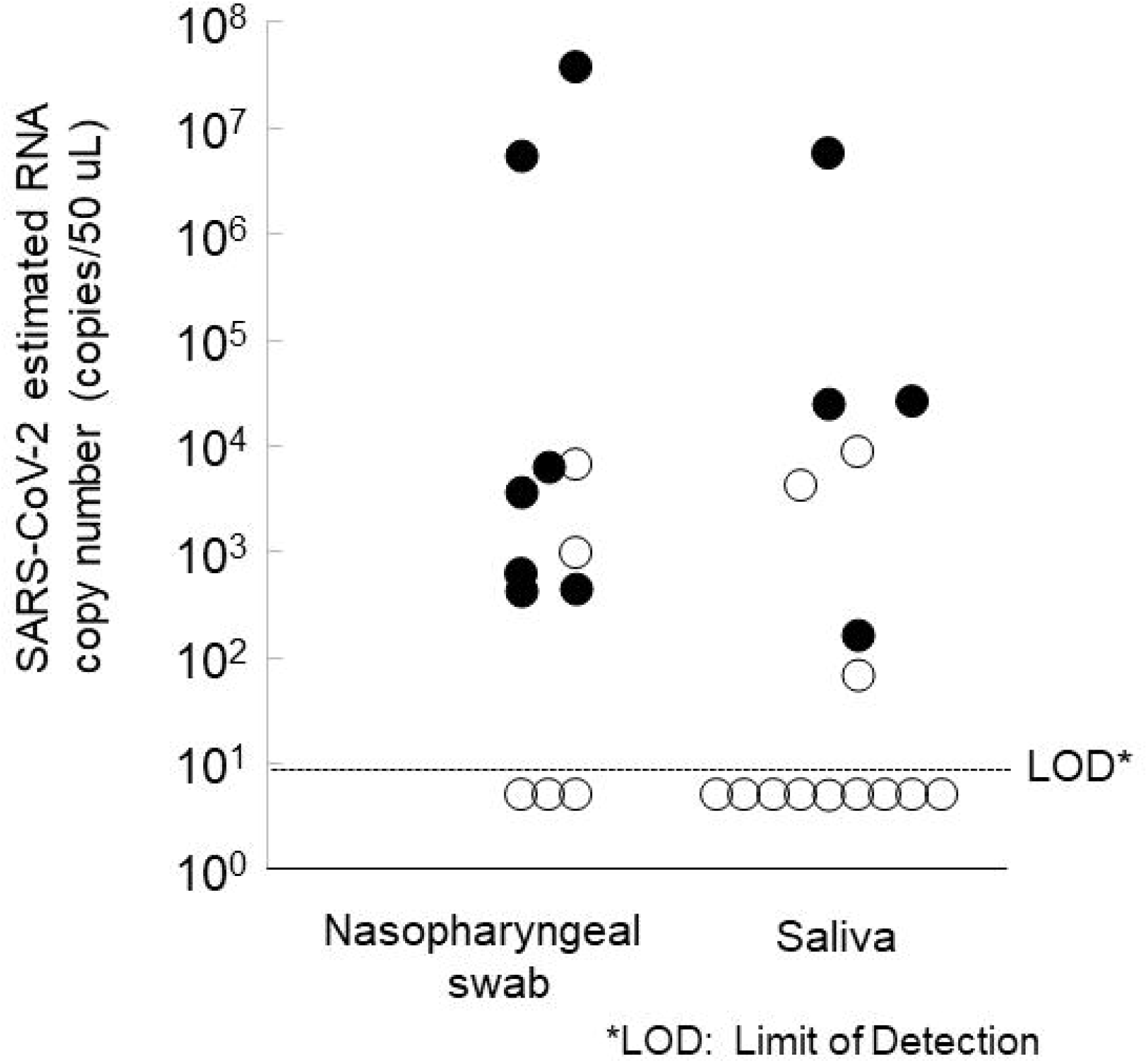
Detection of SARS-CoV-2 N antigen in nasopharyngeal swab and saliva specimens. Closed circles indicate N antigen positive specimens. Among 16 nasopharyngeal swabs, 4 samples without Ag assay data are not plotted.

## Discussion

Sensitivity of SARS-CoV-2 RT-PCR in saliva specimens was lower than those reported previously[1,2,4,6–8]. Among several procedures of saliva collection which have been applied, we chose the passive drool method to reduce the burden of the patients and the secondary risk to healthcare workers. However, collection procedure should be examined for further studies. In addition, symptoms, periods after the onset of symptoms and infection could affect the sensitivity of the RNA detection in saliva. These factors would also affect the sensitivity of antigen detection using the ESPLINE kit.

This study was carried out using specimens collected from hospitalized patients. The mechanism and hazard of secondary infection caused by asymptomatic carriers and super spreaders through biological materials from them have been discussed[9], To assess the secondary infection hazards, it would be important to measure infectious virus particles in saliva and nasal fluid. Low prevalence of RNA in saliva specimens from the hospitalized patients might provide clues to assessment of the secondary infection hazards. To address this issue, mass-screening using POCT tests would be a powerful tool.

This is the first report of SARS-CoV-2 antigen detection in saliva using a POCT antigen test. The combination of saliva specimen and rapid antigen detection test could reduce burden of medical settings by decreasing not only the risk of secondary infection, but also time of diagnosis and cost of the specialized expensive equipment. It could be useful for COVID-19 screening to prevent COVID-19 epidemic by identifying persons who will spread COVID-19 through saliva.

## Data Availability

Data will be available after publication in peered-review journal upon the request.

## Author contributions

YI, SY, KaA and KT contributed to design this study. KK, TMa, TM, and SaY contributed to collect specimens, KoA and YI contributed to conduct and perform RT-PCR and antigen tests, YI, KoA, SY and KaA contributed to analyze data, and SY, YI, KoA and KT contributed to prepare this manuscript.

